# Persistence of SARS-CoV-2 in the first trimester placenta leading to vertical transmission and fetal demise from an asymptomatic mother

**DOI:** 10.1101/2020.08.18.20177121

**Authors:** Prajakta Shende, Pradip Gaikwad, Manisha Gandhewar, Pawankumar Ukey, Anshul Bhide, Vainav Patel, Sharad Bhagat, Vikrant Bhor, Smita Mahale, Rahul Gajbhiye, Deepak Modi

## Abstract

Coronaviruses infect the respiratory tract and are known to survive in these tissues during the clinical course of infection. However, how long can SARS-CoV-2 survive in the tissues is hitherto unknown. Herein, we report a case where the virus is detected in the first trimester placental cytotrophoblast and syncytiotrophoblasts five weeks after the asymptomatic mother cleared the virus from the respiratory tract. This first trimester placental infection was vertically transmitted as the virus was detected in the amniotic fluid and fetal membranes. This congenitally acquired SARS-CoV-2 infection was associated with hydrops and fetal demise. This is the first study providing concrete evidences towards persistent tissue infection of SARS-CoV-2, its congenital transmission in early pregnancy leading to intrauterine fetal death.

## Introduction

SARS-CoV-2 infects the respiratory tract and coronavirus-like particles, SARS-CoV-2 RNA and proteins are detected in the cells from bronchoalveolar lavage or lung tissue from patients with COVID-19^1,2^. Beyond the lung, SARS-CoV-2 is also detected in extra pulmonary sites like the submucosal glands, lymphocytes, subcarinal lymph node, kidney, large intestine, spleen, endothelial cells of various tissues and placenta^2,3^. In experimentally induced COVID-19 infection, SARS-CoV-2 RNA is detected in nasal mucosa, pharynx, trachea, lung tissues, gastrointestinal tract, liver, kidney, paratracheal lymph nodes and spleen^4^. In all these cases, the virus is detected in the tissues during acute viremia or just after recovery (nasopharyngeal swab negative). However, how long can the virus survive in the tissues is hitherto unknown. Although viral shedding in the feces is observed for weeks after clearing pulmonary infection^5–7^, whether this is due to persistent viral infection in the gut tissue or are excretory outcomes of oral ingestion of SARS-CoV-2 (through water) is not clear.

While the modes of human to human transmission of SARS-CoV-2 is well established, controversy exist regarding the transmission of virus from mother to unborn fetuses. SARS-CoV-2 infection is reported in a small proportion of babies born to mother with COVID-19^8,9^, whether this is due to transmission in utero or the infection is acquired during the course of delivery or after both remains unclear. The spike (S) protein of SARS-CoV-2 binds to its cell associated and soluble receptor ACE2 and enters for replication using a plethora of host genes, most of which are expressed in the placental trophoblasts throughout development^10–13^. Indeed, viral RNA and protein are detected in placenta of some SARS-CoV-2 infected mothers^3,14^; whether this infection is transmitted to the baby is yet unclear. A recent report did demonstrate the vertical transmission of the virus due to maternal infection in the third trimester^14^ what are the consequences of maternal viral infection in the first trimester of pregnancy is unknown.

Herein, we report a case of active SARS-CoV-2 viremia in the first trimester placental tissue five weeks after the asymptomatic mother cleared the virus from respiratory tract causing in vertical transmission and fetal demise.

## Case Details

A woman in her late twenties, third gravida with 13 weeks of amenorrhea reported for ANC registration at a tertiary care hospital in Mumbai, India. She had earlier obstetric ultrasonography report showing single live intrauterine gestation with a CRL of 14.6 mm corresponding to a gestational age of 7 weeks 6 days. She had conceived spontaneously with no history of any medical or surgical comorbidities. Her obstetric history includes one living child and one first trimester spontaneous abortion. At 8 weeks of gestation, she had history of contact with symptomatic COVID-19 positive patient. Although she was asymptomatic, her reverse transcriptase polymerase chain reaction (RT-PCR) of nasopharyngeal swab was positive for SARS-CoV-2. She was quarantined for 10 days at a local quarantine/ isolation centre and was discharged with further advice of home quarantine as per the prevailing protocol. She remained asymptomatic throughout this period.

On a booking visit for antenatal care five weeks later, she was advised routine blood investigations along with ultrasonography for nuchal translucency. The ultrasound evaluation showed fetal demise with changes of hydrops fetalis. Since there was fetal demise the products of conception were sent for further routine evaluation. SARS-CoV-2 testing was done out of scientific curiosity.

## Methods

The study was approved by the Institutional Ethics Committee of ESI- PGIMSR and Model Hospital, Mumbai and ICMR-NIRRH, Mumbai India. Written informed consent was taken. Dilation and curettage was done according to hospital norms. All necessary precautions were taken to avoid contamination of the placental and amniotic fluid samples. Serum was sent for TORCH testing. Piece of the placental tissues were fixed in 10% buffered formalin processed for paraffin embedding and sectioning. Placental histopathology was evaluated by routine Hematoxylin and Eosin staining. Fluorescence *in situ* hybridization on paraffin sections was done to determine aneuploidy of chromosomes 13, 18, 21, X and Y. Placental villi were collected in virus transport medium. Amniotic fluid was carefully aspirated from the gestational sac and transported in a sterile container. RT-PCR for SARS-CoV-2 were done as per the ICMR/WHO Guidelines for RdRP and E genes. Immunofluorescence for SARS-CoV-2 spike proteins S1 and S2 on placental sections was done using monoclonal antibodies (MP Biomedicals, Catalogue number: SKU: 08720301, SKU: 08720411, Asia Pacific, Singapore) and the Opal 7 (catalogue #NEL797001KT; PerkinElmer, Waltham, MA), which uses individual tyramide signal amplification (TSA)-conjugated fluorophores.

## Results

### First trimester SARS-CoV-2 infection is associated with hydrops fetalis and fetal demise

At 8 weeks of amenorrhea, ultrasonography revealed a single live fetus with a CRL of 14.6 mm corresponding to 7 weeks 6 days of gestation. She was asymptomatic at presentation; her throat swab was positive for ORF1b gene of SARS-CoV-2 with a Ct Value of 28.

The repeat throat swab was taken the day prior to termination (six weeks after the first positive report) which was negative for SARS-CoV-2. Subsequently the ultrasound evaluation (expectedly 13 weeks of gestation) showed fetal demise at crown rump length (CRL) of 38mm corresponding with 10 weeks 5 days of gestation. There was extensive bilateral pleural effusion and subcutaneous edema suggestive of hydrops fetalis (Fig.1).

**Fig 1.**
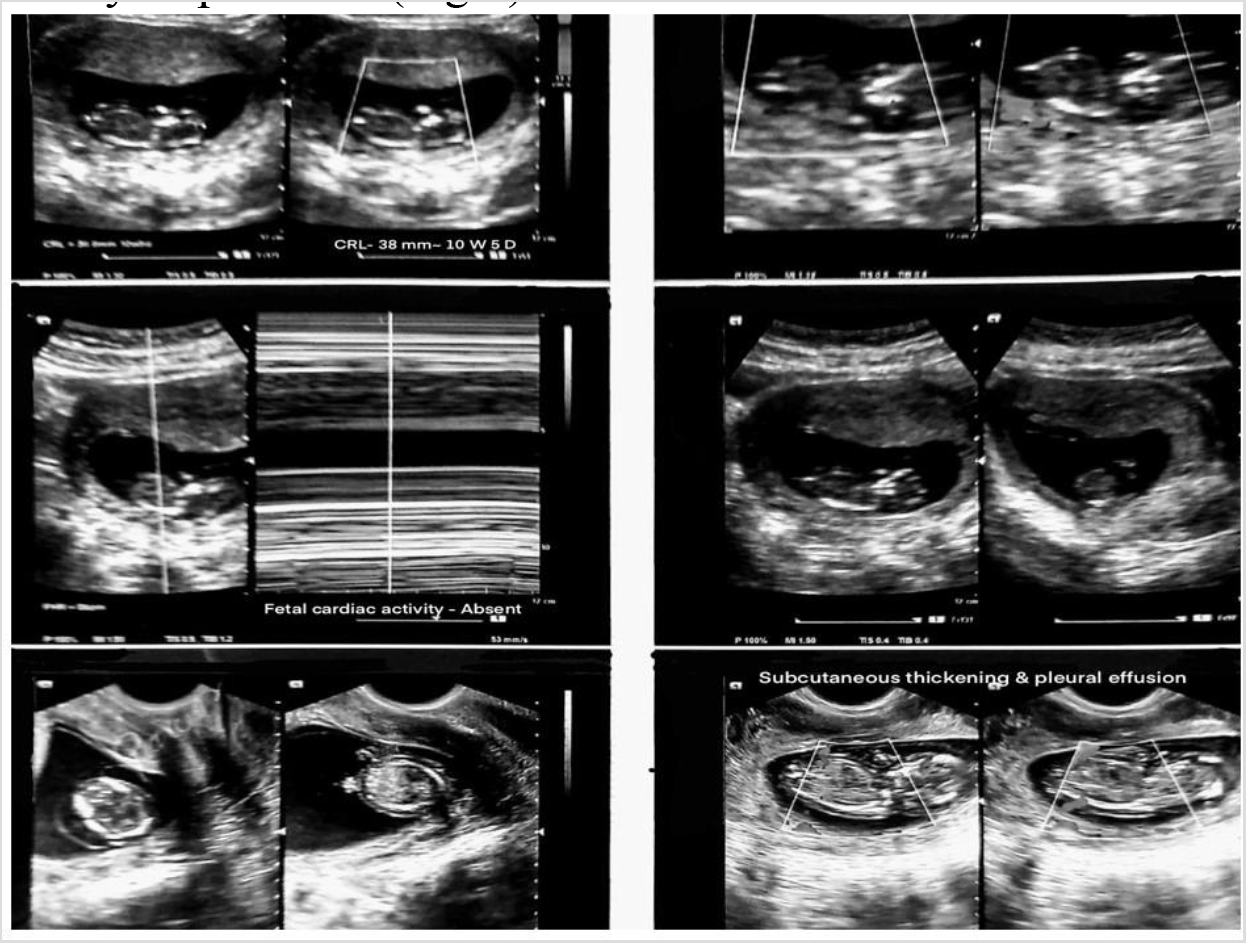
Ultrasonography images of the 13 weeks gestation showing fetal demise with changes of hydrops fetalis.

She was negative for antiphospholipid antibodies, anti-nuclear antibodies and anticardiolipin antibodies ruling out this as possible cause of first trimester abortion. IgM antibodies against TORCH were negative which indicates absence of active infection, however IgG antibodies for Toxoplasma gondii, Rubella, Cytomegalovirus and Herpes simplex virus 1 were above the reference range. (**Supplementary Table 1**). Her blood group was O+ve ruling out possibility of autoimmune cause of hydrops. Fluorescence *in situ* hybridization of placental villi showed disomy for 13, 18, 21, and X chromosomes ruling out possibility of major chromosome aneuploidies causing hydrops fetalis (not shown).

### Persistent presence of SARS-CoV-2 in placental cells

RNA for both E gene and RdRp gene of SARS-CoV-2 were detected in the supernatants with a Ct value of 28.4 and 27.5 respectively suggestive of moderate viral load (Table 1). To determine if SARS-CoV-2 has infected placental cells we carried out immunofluorescence (Fig 2) using monoclonal antibodies against the spike protein S1 and S2 on placental sections. Both, S1 and S2 proteins were diffusely localized mainly in the cytoplasm of syncytiotrophoblasts and some cytotrophoblasts cells. Some villus stromal cells were also positive for both the proteins. In some syncytiotrophoblasts, S2 was also found as aggregates. These results indicate that the viremia persisted in the placenta six weeks after the mother was tested positive.

**Table 1:** Result of RT-PCR for SARS-CoV-2 in the nasopharyngeal swabs, placenta and amniotic fluid from a pregnant woman with COVID-19 in first trimester. Maternal nasopharyngeal swab I collected at 8 weeks of gestation (during active infection). Maternal nasopharyngeal swab II is collected at 13 weeks of gestation (5 weeks after active infection). Viral carriage is placenta was estimated by incubating placental villi in virus transport medium. Viral carriage in amniotic fluid was estimated in the fluid aspirate of the gestational sac. Ct values are cycle threshold value for each sample.

| Tissue | Gene tested | Ct value | Result |
| --- | --- | --- | --- |
| Viral carriage in maternal Nasopharyngeal swab I | Orfb1 | 28 | Positive |
| Viral carriage in maternal Nasopharyngeal swab II | Orfb1 | 0 | Negative |
| Viral carriage in placenta | E gene | 28.4 | Positive |
|  | RdRp | 27.5 |  |
| Viral carriage in amniotic fluid | E gene | 26.3 | Positive |
|  | RdRp | 25.4 |  |
| Negative control | E gene | ND | Negative |
|  | RdRp | ND |  |

**Fig 2.**
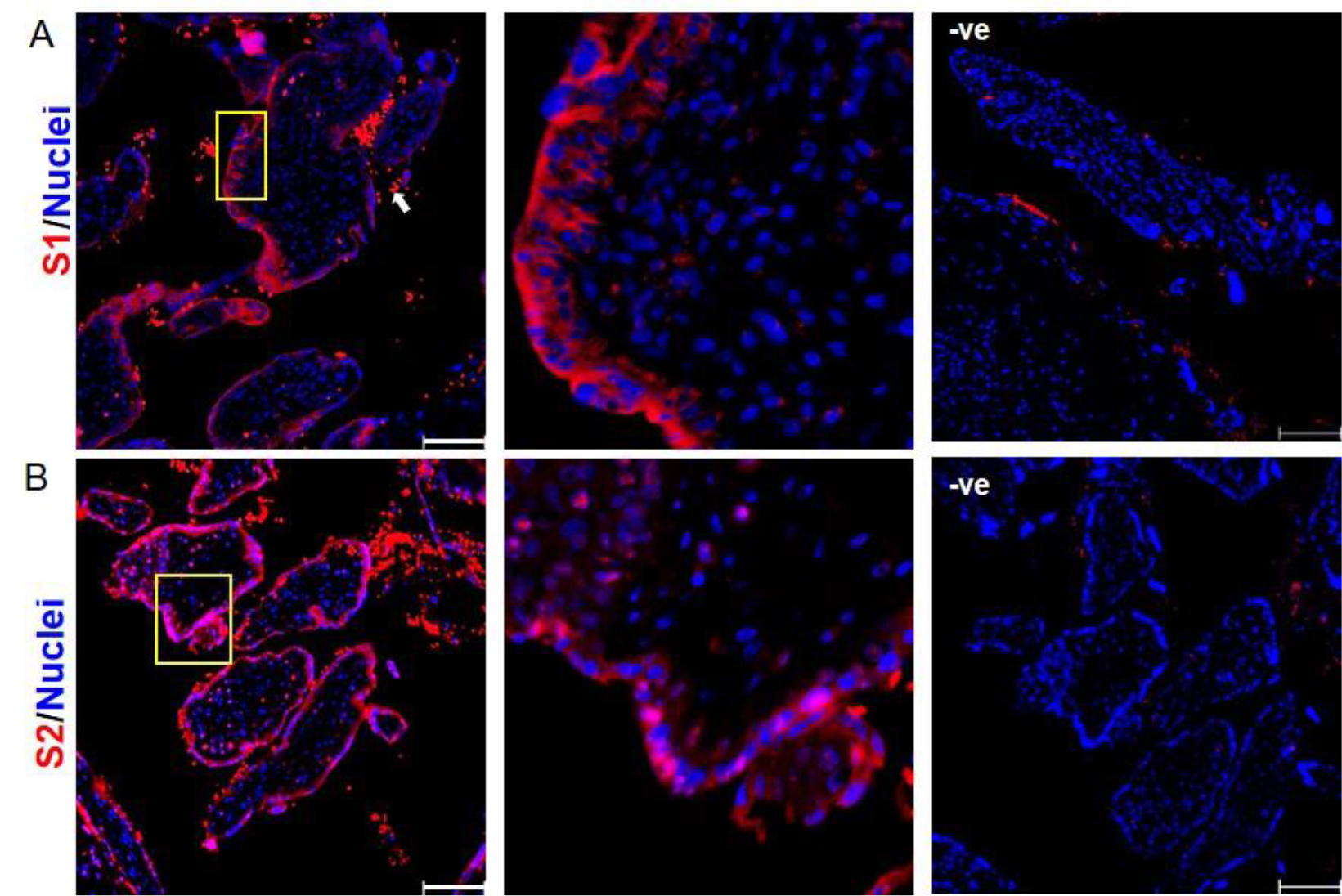
Detection of Spike Proteins of SARS-CoV-2 in villi of first trimester placenta of a women with COVID-19. Paraffin sections were immunostained for Spike proteins S1 (Panel A) and S2 (Panel B) of SARS-CoV-2 using monoclonal antibodies. Boxed area is enlarged in the next panel to show the specific cell types. Negative are sections of same tissues incubated without primary antibody. In all the sections the read staining in the inter villus spaces is autofluorescence of red blood cells. Scale bar represents 100 μm.

### Histopathology of the placenta

Placental histopathology (Fig 3) revealed conspicuously avascular villi with extensive perivascular fibrin deposition. The intravillous area had large numbers of stromal cells and with extensive cytoplasmic vacuolation. In some areas the syncytiotrophoblast layer appeared lysed. The decidua also had extensive fibrin deposition and large dilated blood vessels engorged with RBCs. There were signs of extensive inflammation as evident by presence of large numbers of leucocytes including the polymorphonuclear leucocytes in the decidual bed as well as in the intervillous spaces.

**Fig 3.**
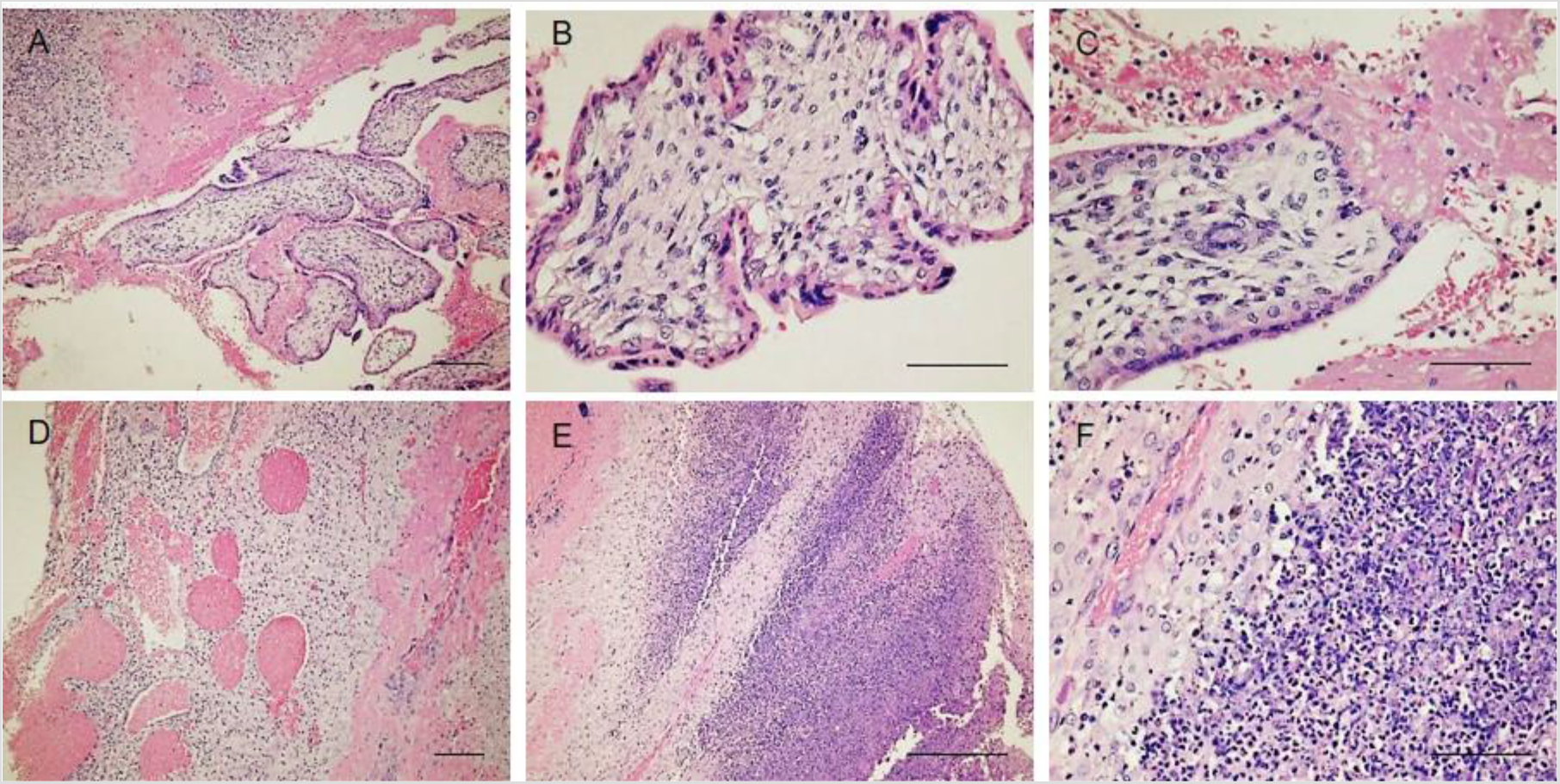
Histopathology of placenta and decidual tissue from a woman with SARS-COV2 in first trimester. Panel A-C are placental villi and D–F is the decidual tissue stained with haematoxylin and eosin. A) is lower magnification image showing fibrin deposition in the intervillous space and decidua. B) shows vacuolated avascular villi, C) shows lysis of syncytiotrophoblast cells in the villi, leucocyte infiltration in intervillous spaces and fibrin deposition. D is a low magnification image showing shows dilated blood vessels in the decidua and fibrin deposition, E shows infiltration of immune cells in the decidual tissues and F) is higher magnification of E showing leucocytes in the decidual bed. In all the images scale bar represents 100 μm

### Vertical transmission of SARS-CoV-2

To test if placental SARS-CoV-2 is vertically transmitted, we carried out RT-PCR in the amniotic fluid and RNA for both E and RdRp genes were found at Ct value of 26.3 and 25.4 respectively (Table 1). The viral S proteins were also detected in cells of fetal membrane. Both S1 and S2 were diffusely localized in cytoplasm of cells of placental membranes (Fig 4).

**Fig 4:**
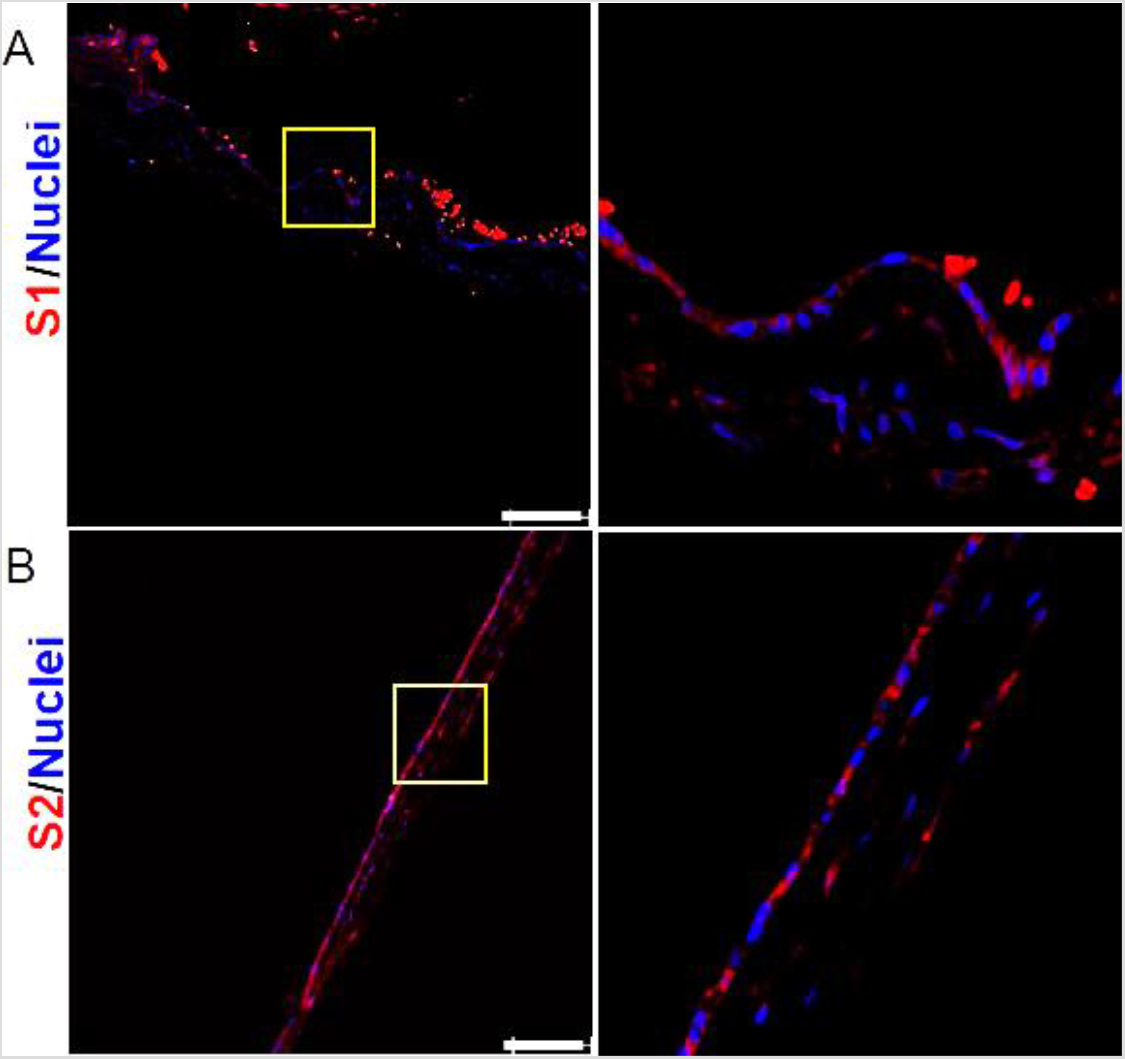
Detection of Spike Proteins of SARS-CoV-2 in first trimester fetal membrane from a women with COVID-19 in first trimester. Paraffin sections were immunostained for Spike proteins S1 (Panel A) and S2 (Panel B) of SARS-CoV-2 using monoclonal antibodies. Boxed area is enlarged in the next panel to show the specific cell types. Scale bar represents 100 μm.

## Discussion

To our knowledge, this is the first case demonstrating persistence of SARS-CoV-2 infection in a tissue weeks after clearance in the throat swabs. The study also provides evidence for vertical transmission of the virus in the first trimester leading to fetal demise.

Prior studies have shown persistent viral shedding in feces of COVID-19 patients, the source of this virus is unknown. Herein, we show that while the mother was negative for SARS-CoV-2 in the pulmonary passage, extensive viremia was observed in the placenta almost five weeks of the initial infection. Both “E” and “RdRp” gene of SARS-CoV-2 were found in the placental supernatants at viral loads equivalent of that of the maternal nasopharyngeal swab observed earlier. Thus, the virus not just survived in the tissue but is replicative in the placental cells. Along with viral RNA, the viral proteins could be readily detected in the cytotrophoblasts, syncytiotrophoblasts and some villus stromal cells. Indeed, we have shown that all these cell types in first trimester placenta not only expresses the canonical (ACE2) and the non-canonical (CD147) SARS-CoV-2 receptors, they readily express the proteins required for viral endocytosis, replication and release^11^. Thus, the placenta is a permissive site for SARS-CoV-2 replication and such long-term persistence of the virus in any tissues post clearance from the pulmonary sites has not been shown previously.

The active viremia of the placental cells was further coupled with extensive placental inflammation as deduced by leucocyte infiltration in the intravillous spaces and decidua, fibrin deposition and lysis of syncytiotrophoblasts of the villus cells. Inflammatory changes and fetal vascular malperfusion or fetal vascular thrombosis are reported in second and term placental tissues obtained from mothers infected with COVID-19 and active viremia^3,14–16^. These observations imply that SARS-CoV-2 not just reside and replicate in placental cells but also mount an inflammatory response.

Mother to child transmission of viruses can cause congenital anomalies and is a matter of concern in SARS-CoV-2 pandemic. Epidemiological evidence suggests a low possibility of mother to child transmission of SARS-CoV-2; however, this notion is based on reports of third trimester infections^8,9^. Also, most reports are classified only as probable case of congenital SARS-CoV-2 infection as the infection of the newborn is reported hours or even days after birth^9^. Our case fully qualifies as congenitally transmitted SARS-CoV-2 infection as we not only detected the virus in the placental cells but also in the amniotic fluid and the fetal membrane. Recently, a case of transplacental transmission of SARS-CoV-2 in a neonate born to a mother infected at term was reported^14^. While SARS-CoV-2 is not a blood borne virus, it is possible that some SARS-CoV-2 particles may have seeped in to the maternal blood stream or transported through the lymphocytes during the course of active infection, resided and replicated in the placenta and eventually crossed the transplacental barrier to infect the fetus.

A variety of viral infections, chromosome abnormalities and autoimmune conditions are associated with nonimmune hydrops fetalis^17,18^. In our case, although the mother was positive for IgG of herpes simplex virus and Toxoplasma gondii, she was negative for IgM of the same ruling out active infection with these organisms as a cause of hydrops. Also, the mother was Rh positive and product of conception was disomic for chromosomes 13, 21, 18 and X ruling out these factors in etiology of hydrops. We believe that the hydrops and fetal demise are due to congenital transmission of SARS-CoV-2 from the mother to the fetus. To the best of our knowledge this is the first study associating hydrops fetalis and fetal demise with congenitally acquired coronavirus.

## Conclusion

This is the first study providing concrete evidences towards persistent tissue infection of SARS-CoV-2, its congenital transmission in early pregnancy leading to hydrops fetalis and fetal demise. Further studies are required to throw more light on vertical transmission of SARS-CoV-2 infection in first trimester so that universal screening of all pregnant women can be considered in such cases to avoid adverse fetal outcome.

## Data Availability

All the data is available from the authors upon request

## Acknowledgements

The manuscript bears NIRRH ID RA/956/08–2020 and is a part of National Registry of Pregnant Women with COVID-19 in India (PregCovid Registry) https://pregcovid.com/. The authors declare no conflict of interest.

---

**Supplementary Table 1:** Outcomes of the TORCH test in maternal serum at 13 weeks of gestation of a woman with SARS-CoV-2 at 8 weeks of gestation.

| APLA /Anti DNA /TORCH<br>Antibodies | Value | Reference Range |  |  |
| --- | --- | --- | --- | --- |
|  |  | Negative | Equivocal | Positive |
| Antiphospholipid (APLA) IgG | 2.83 | < 12 | 12 -18 | >18 |
| Antiphospholipid (APLA) IgM | 4.09 | <12 | 12 -18 | >18 |
| Anti-double stranded DNA | 0.34 | < 0.90 | 0.90 -1.10 | > 1.10 |
| Cardiolipin (ACL) IgG | 3.89 | < 12 | 12-18 | >18 |
| Cardiolipin (ACL) IgM | 5.7 | < 12 | 12-18 | >18 |
| Cytomegalovirus (CMV) IgG | 4.26 | < 0.80 | 0.80 - 1.20 | >1.20 |
| Cytomegalovirus (CMV) IgM | 0.47 | <0.80 | 0.80 – 1.20 | >1.20 |
| Toxoplasma gondii IgG | 6.52 | <0.70 | 0.70 – 1.30 | >1.30 |
| Toxoplasma gondii IgM | 0.46 | <0.80 | 0.80 – 1.20 | >1.20 |
| Rubella IgG | 3.16 | <0.70 | 0.70 – 1.30 | >1.30 |
| Rubella IgM | 0.52 | <0.80 | 0.80 – 1.20 | >1.20 |
| Herpes simplex virus (HSV) I - IgG | 5.07 | <0.80 | 0.80 – 1.20 | >1.20 |
| Herpes simplex virus (HSV) II - IgG | 0.27 | <0.80 | 0.80 – 1.20 | >1.20 |
| Herpes simplex virus (HSV) I - IgM | 0.59 | <0.80 | 0.80 – 1.20 | >1.20 |
| Herpes simplex virus (HSV)II - IgM | 0.46 | <0.80 | 0.80 – 1.20 | >1.20 |
| Beta 2 Microglobulin | 1916 | 670-2143 |  |  |

